# Effectiveness of Pasteurization for the Inactivation of H5N1 Influenza Virus in Raw Whole Milk

**DOI:** 10.1101/2024.07.23.24310825

**Authors:** Tamiru Alkie, Neda Nasheri, Pablo Romero-Barrios, Angela Catford, Jay Krishnan, Lemarie Pama, Kathleen Hooper-McGrevy, Charles Nfon, Todd Cutts, Yohannes Berhane

## Abstract

Highly pathogenic avian influenza (HPAI) clade 2.3.4.4b H5Nx viruses continue to cause episodic incursions and have been detected in more than 12 taxonomic orders encompassing more than 80 avian species, land and marine mammals, including recent detections in dairy cattle. The HPAI H5N1 spillover to these important livestock species creates a new interface for human exposure and raises food safety concerns. Presence of H5N1 genetic material in one out of five retail pasteurized milk samples in the USA has prompted the evaluation of pasteurization processes for the inactivation of influenza viruses. Our study examined whether pasteurization could effectively inactivate HPAI H5N1 inoculated raw whole milk samples. We heated 1 mL of non-homogenized cow’s milk samples to attain an internal temperature of 63°C or 72°C and spiked with 6.3 log EID_50_ of clade 2.3.4.4b H5N1 virus. Complete inactivation was achieved after incubation of the H5N1 spiked raw milk at 63°C for 30 minutes. In addition, complete viral inactivation was observed in seven out of eight replicates of raw milk samples treated at 72°C for 15 seconds. In one replicate, a 4.56 log reduction was achieved, which is about 1 log higher than the average viral quantities detected in bulk tanks in affected areas. Therefore, we conclude that pasteurization of milk is an effective strategy for mitigation of risk of human exposure to milk contaminated with H5N1 virus.

## 1. Introduction

The spread of HPAI H5Nx viruses bearing clade 2.3.4.4b hemagglutinin (HA) and their variants generated by reassortment with low pathogenicity avian influenza viruses (LPAIVs) remain the major causes of large-scale HPAI outbreaks in domestic poultry and wild birds worldwide (Youk, et al. 2023, Alkie, et al. 2022). They have also caused multiple spillover infections into mammals resulting in mass die-offs in the Americas (Rimondi, et al. 2024, Alkie, et al. 2023). However, naturally occurring HPAI infections in ruminant livestock have never been reported before 2024. The fast spread of HPAI virus in dairy cows, as well as sustained transmission among dairy herds in 13 states in the USA since March 2024 indicates wider host ranges of HPAI clade 2.3.4.4b H5N1 viruses (FDA.). The detection of infectious virus in milk samples obtained from infected dairy cows indicate the potential risk of accidental introduction of H5N1 virus into the human food supply. Although human foodborne infection with H5N1 virus has not been confirmed to date, exposure to contaminated milk in mice and cats resulted in severe and fatal infections (Guan, et al. 2024, Burrough, et al. 2024).

Dairy milk is usually sold pasteurized, and the sale of raw milk is not permitted in some jurisdictions like Canada (Health Canada.). Based on the understanding of viral structure and previous studies on pasteurization of eggs and human milk (Chmielewski, et al. 2011, Swayne and Beck. 2004, Pitino, et al. 2021), it is assumed that pasteurization of milk would be effective against H5N1 viruses, but recent studies have demonstrated a range of results (Cui, et al. 2024, Schafers, et al. 2024, Kaiser, et al. 2024, Guan, et al. 2024). Also, studies performed using other viruses have demonstrated that certain milk components such as fat globules and casein micelles may protect viruses against heat treatment (Tomasula, et al. 2007). Although traces of H5N1 genomic materials have been detected in about 20% of retail dairy samples that were sampled in USA supermarkets, infectious H5N1 viruses have never been isolated from retailed pasteurized dairy products to date (FDA.). With the ongoing rapid virus evolution and spread of H5N1, more dairy farms could be infected in the USA and potentially in other countries. Should raw milk from infected asymptomatic cows accidentally enter the processing chain for human consumption, pasteurization can be an effective treatment to inactivate the H5N1 virus. In the current study, we simulated pasteurization processes typically used by the dairy industry on unpasteurized and unhomogenized raw milk inoculated with H5N1 clade 2.3.4.4b at titers that have been reported in the affected farms (Spackman, et al. 2024).

## 2. Materials and Methods

### 2.1. Heat inactivation at 63°C for 30 minutes

The temperature combination of 63°C for 30 minutes represents a vat or bulk pasteurization treatment sometimes used in commercial dairy processing plants. Firstly, fresh bulk raw milk obtained from a dairy farm in Manitoba, Canada was treated for 1 hr at room temperature with a cocktail containing antibiotics and antifungals (streptomycin, penicillin, vancomycin, nystatin and amphotericin) (Sigma-Aldrich) to prevent any harmful effects of bacteria on the embryonated chicken eggs used to test the infectiousness of HPAI H5N1 virus. The sterility of the milk samples from bacteria was confirmed by inoculating into Columbia Blood Agar Base medium and monitored daily for bacterial growth. The entire procedure was conducted in a biosafety level 3+ (BSL-3+) facility at the National Centre for Foreign Animal Disease (NCFAD) in Winnipeg, Manitoba. Nine-hundred and ninety microliters milk (990 µL) each was placed in four 1.5 mL Eppendorf tubes and was heated in a water bath to attain an internal temperature of 63°C. The temperature was monitored using the Applied Biosystems Thermal Cycler Temperature Verification System (Model 4500). The raw milk in all four tubes was spiked with 10 µL of clade 2.3.4.4b H5N1 virus, recapped and treated for 30 minutes. The virus was designated as A/Turkey Vulture/Ontario/FAV473-3/2022 having a stock titer of 1 × 10^8.3^EID_50_/mL isolated from Canada and archived at NCFAD. The final virus concentration before heat inactivation was calculated at 6.3 log_10_EID_50_. Tubes were then placed on ice to cool down their contents. The contents of three of the four tubes were inoculated through the allantoic route into 9-days-old embryonated specific-pathogen-free (SPF) chicken eggs (ECEs) following the standard protocol described in the Manual of Diagnostic Tests and Vaccines for Terrestrial Animals (World Organisation for Animal Health. 2023)(n = 5 eggs/tube and 200 µL per egg) and incubated at 37°C. The fourth tube (replicate 4) was serially diluted (10-fold) and inoculated into ECEs (n = 5 eggs/dilution) to determine the log reduction of the virus following treatment at 63°C for 30 minutes. Allantoic fluid was collected from each dead or live embryo during the first and second passages in ECEs and checked by hemagglutination assay using the standard protocol described above (World Organisation for Animal Health. 2023) to confirm the presence of infectious avian influenza viruses. The temperature of three non-spiked milk samples heated at 63°C and kept on ice were recorded every minute until reaching 4°C.

### 2.2. Heat inactivation at 72°C for 15 seconds

The temperature combination of 72°C for 15 seconds represents a high temperature short time (HTST) pasteurization treatment commonly used in commercial dairy processing plants. This heat inactivation experiment was conducted in two independent trials conducted on different days. The raw milk was treated with the antibiotic cocktails as described above. The heating of raw milk to attain an internal temperature 72°C was conducted in a heating block (Eppendorf Thermomixer model R) and the temperature was monitored using an electronic temperature probe inserted into two tubes containing raw milk samples (temperature control) when the first and last experimental samples were spiked and heat-treated. The heating block temperature was increased to 76°C to attain internal milk temperature of 72°C based on the Applied Biosystems Thermal Cycler Temperature Verification System reading. Ten microliters (10 µL) of virus were spiked into the raw milk when the internal temperature of the milk reached 72°C and after 15 seconds of incubation, the lid of the Eppendorf tube was recapped and placed on ice to cool. This procedure was performed to remove possible impact of the temperature ramp up, which would not be experienced in a commercial pasteurization process where nearly instantaneous changes in temperature are experienced by fluid milk passing through small diameter tubing or pipe. The procedure of spiking of the virus into the raw milk, amount and concentration of virus and ECE inoculation procedures were the same for the 63°C for 30 minutes treatment protocol. All four tubes in each trial were then tested in embryonated ECE as described above.

The gradual temperature decline on ice was recorded in three non-spiked milk samples treated as mentioned above.

## 3. Results

The allantoic fluids from dead and live chicken embryos were collected separately (without pooling) at the end of the 1^st^ and 2^nd^ passages and were individually checked for HA activity. Complete H5N1 virus inactivation was achieved in all milk samples (four replicates) treated at 63°C for 30 minutes. No HA activity was detected in any of the allantoic fluid harvested from live embryonated eggs during the first and second passages. Moreover, allantoic fluid collected from embryonated eggs that died of non-specific causes (likely from injections of 200 µL milk) was negative for any HA activity. In addition, complete viral inactivation was achieved in 7 out of 8 replicates treated at 72°C for 15 seconds and no infectious virus was recovered in ECE in either passage. A 4.56 log reduction in virus titers was achieved in the eighth replicate. We observed that milk treated at 72°C for 15 seconds underwent progressive temperature decline (Figure 1) when kept on ice, mimicking the pasteurization and chilling processes in the processing plants. Within one minute on ice, milk treated at 72°C dropped to 56°C and after 5 minutes, the temperature reached 25°C (Figure 1). A similar temperature decline trend was observed for the milk treated at 63°C. Therefore, we can conclude that pasteurization of milk at the two standard heat treatments used by industry can effectively inactivate clade 2.3.4.4b H5N1 viruses in raw milk that may have incoming viral titres of approximately 4.5 and 6 log_10_ EID_50_.

**Figure 1.**
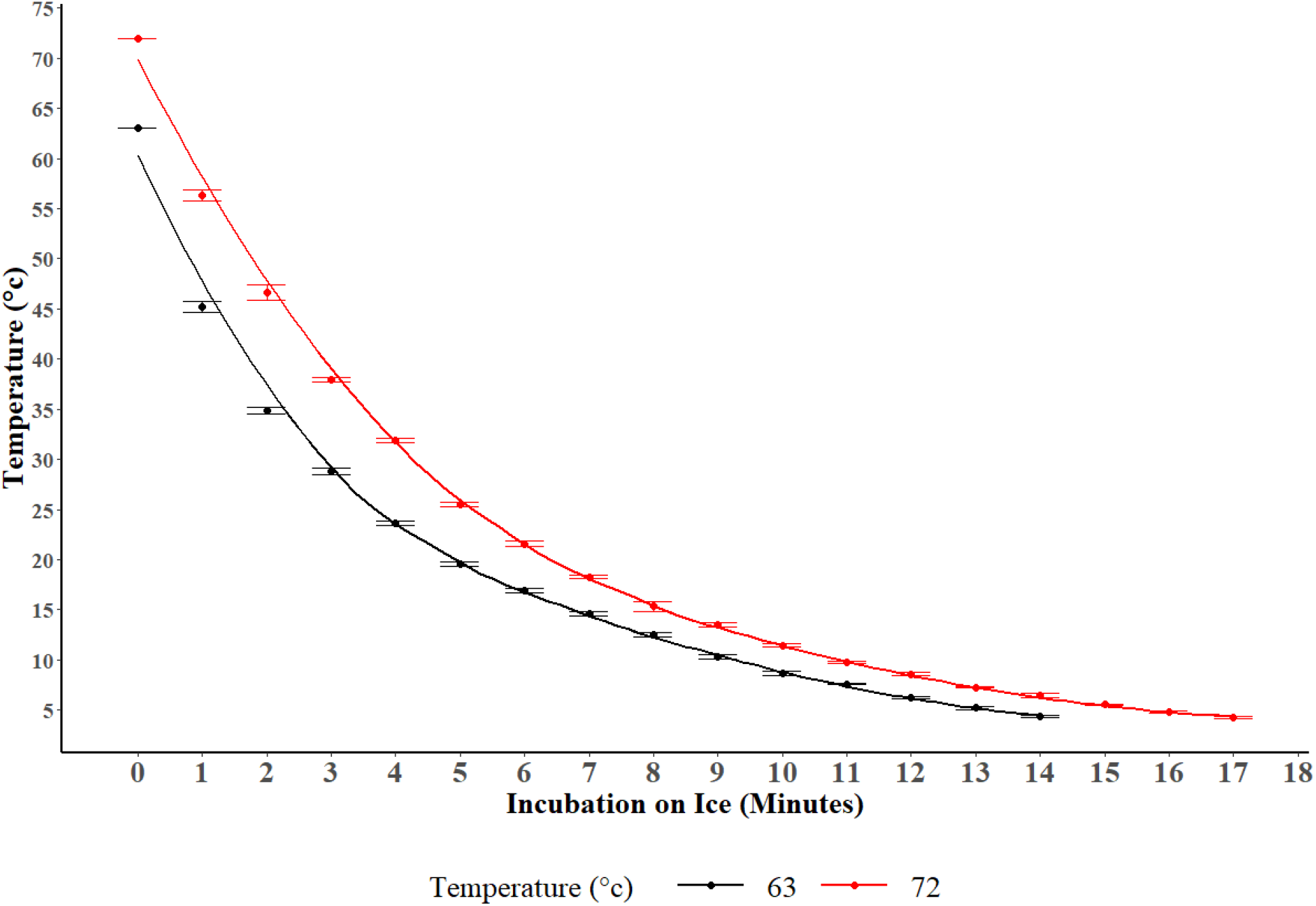
Temperature plot of cooling whole milk after heat treatment and incubation on crushed ice. The dots represent the average temperature of three replicates and error bars represent standard error. The pasteurization at 63°C for 30 minutes is shown in black color and the treatment at 72°C for 15 seconds is shown in red.

## 4. Discussion

The spillover and transmission of HPAI A(H5N1) clade 2.3.4.4b virus to dairy cows in the USA and the regular detection of infectious virus in raw milk obtained from infected cows point to the potential risk of human exposure to the virus as a result of the handling or ingestion of raw milk and dairy products. Since spring of 2024, various regulatory agencies in the US and Canada have tested retailed pasteurized milk samples for the presence of HPAI H5N1 viruses and evaluated the associated health risks. Studies in the USA have shown the presence of H5N1 virus genetic fragments often in significant concentrations in pasteurized retail milk samples, however, follow up tests failed to detect any infectious H5N1 virus (FDA.). This prompted public health officials to declare that consumption of pasteurized retail milk was safe (FDA.).

While the knowledge of general viral behaviour for influenza A viruses, and previous data (Chmielewski, et al. 2011, Swayne and Beck. 2004, Pitino, et al. 2021) allows for a sound extrapolation that pasteurization would likely be effective to inactivate HPAI clade 2.3.4.4b H5N1 virus in milk from dairy cows, the detection of this virus in this species and matrix is a new phenomenon. As we did not have access to any milk or milk products from dairy cows naturally infected with HPAI virus, we conducted an independent study by inoculating H5N1 virus into milk samples to evaluate the efficiency of standard pasteurization treatments in Canada in inactivation of H5N1 virus. Although we used a benchtop method for heat inactivation, the pasteurization, and cooling steps performed simulate the commercial pasteurization process, which was tested in the recent H5N1 inactivation study using a modified continuous flow pasteurizer method (Spackman, et al.)

We observed that heat treatment at 63°C for 30 minutes effectively inactivates H5N1 virus present in the milk, as supported by all recent published literature (Cui, et al. 2024, Schafers, et al. 2024, Kaiser, et al. 2024, Guan, et al. 2024). The combination of inoculation of ECEs and confirmation of live viruses using the HA assay is considered the most sensitive method of detection for this virus, compared to cell culture assays.

Thus, we can conclude that the standard vat pasteurization at 63°C for 30 minutes guarantees virus inactivation and food safety.

Heat treatment of milk samples inoculated with H5N1 viruses at 72°C for 15 seconds resulted in complete virus inactivation in 7 out of 8 replicates and a significant reduction of infectious virus in the remaining replicate. This finding is consistent with recent reports where residual levels of infective viruses were detected in naturally contaminated milk upon treatment at 72°C for 15 seconds (Guan, et al. 2024, Kaiser, et al. 2024). Two other recent studies assessing the effectiveness of milk pasteurization to inactivate influenza virus found that treatment at 72°C for 15 seconds was effective in fully eliminating infectivity below the limit of detection (Schafers, et al. 2024, Cui, et al. 2024). In one of the two studies, the total volume of inoculated milk for each treatment was only 10 µL, which is a small volume compared to the present study (Cui, et al. 2024). Together, these existing published reports of heat inactivation of influenza viruses in milk employed PCR thermocyclers and relatively smaller milk volume.

A final point of comparison relates to the speed of temperature changes for treated samples in order to best mimic real-life scenarios. In the PCR thermocyclers heating and cooling setup, milk treated at 72°C can instantly cool within seconds (Cui, et al. 2024). In our heat treatment and chilling protocols, we sought to mimic the refrigeration processes of pasteurized milk in dairy processing plants, creating a gradual decline in milk temperature during refrigeration which may still inactivate residual H5N1 viruses. In the modified continuous flow pasteurizer study to examine the effectiveness of HTST inactivation method, the H5N1 inoculated milk was preheated at 40°C. An additional heating for 9 seconds at 72°C before a HTST pasteurization could have an additive effect on virus inactivation (Spackman, et al. 2024). Herein we observed complete inactivation of H5N1 virus in 7 out of 8 replicates at 72°C for 15 seconds, and 4.56 log reduction in one of the replicates, which is one log greater than the mean quantity of infectious virus detected in raw milk from bulk storage tank of naturally contaminated samples (Spackman, et al. 2024).

The experimental studies conducted by inoculating known concentrations of viruses into milk do not fully reflect the actual commercial milk pasteurization and refrigeration processes, yet experiments which simulate pasteurization and gradual chilling steps do provide valuable evidence that H5N1 viruses can be effectively inactivated. Although there is no evidence of foodborne transmission of HPAI H5N1 virus to humans, our data suggest that the pasteurization of milk could effectively mitigate the risk of human exposure to live virus, and subsequent infections should the foodborne route be proven. Also, a longer treatment at low temperature scheme is a more effective method of H5N1 inactivation in dairy products during unprecedented large scale HPAI outbreaks in dairy cows.

## Data Availability

All data produced in the present study are available upon reasonable request to the authors

## Acknowledgements

This study is financially supported by the Defence Research and Development Canada (DRDC) CW2248375 -39903-23076 CSSP 2541 and CFIA HPAI Emergency Grant as well as Health Canada. The authors would like to thank Dr. Sean Li and Dr. Sandeep Tamber for reviewing the manuscript and providing insightful comments.

